# *Plasmodium falciparum* populations, transmission dynamics and infection origins across Papua New Guinea

**DOI:** 10.1101/2023.09.04.23294444

**Authors:** G.L. Abby Harrison, Somya Mehra, Zahra Razook, Natacha Tessier, Stuart Lee, Manuel W. Hetzel, Livingstone Tavul, Moses Laman, Leo Makita, Roberto Amato, Olivo Miotto, Nicholas Burke, Anne Jensen, Dominic Kwiatkowski, Inoni Betuela, Peter M. Siba, Melanie Bahlo, Ivo Mueller, Alyssa E. Barry

**Affiliations:** Division of Population Health and Immunity, Walter and Eliza Hall Institute of Medical Research, Parkville, Victoria, AUSTRALIA; Department of Medical Biology, University of Melbourne, Carlton, Victoria, AUSTRALIA; Burnet Institute, Melbourne, AUSTRALIA; IMPACT/School of Medicine, Deakin University, Geelong, AUSTRALIA; Swiss Tropical and Public Health Institute, Basel, SWITZERLAND; University of Basel, Basel, SWITZERLAND; Papua New Guinea Institute of Medical Research, Madang, PAPUA NEW GUINEA; National Department of Health, Port Moresby, PAPUA NEW GUINEA; Wellcome Sanger Institute, Hinxton, UNITED KINGDOM; University of Oxford, Oxford, UNITED KINGDOM; Mahidol-Oxford Research Unit, Mahidol University, Bangkok, THAILAND; Exxon Mobil PNG Ltd, Port Moresby, PAPUA NEW GUINEA; Divine Word University, Madang, PAPUA NEW GUINEA; Parasites and Insect Vectors, Institut Pasteur, Paris, FRANCE

**Keywords:** malaria, *Plasmodium falciparum*, genomics, single nucleotide polymorphisms, diversity, population genetics, spatial epidemiology, control

## Abstract

Pathogen genomic surveillance demands rapid, low-cost genotyping solutions for tracking infections. Here we use single nucleotide polymorphism (SNP) barcodes to generate practical information for malaria surveillance and control. Using 91 *Plasmodium falciparum* genomes from three provinces of Papua New Guinea (PNG), we assessed SNP panels with different allele frequency characteristics. A 191 ‘local’ SNP barcode captured similar patterns of population structure evident with 5786 ‘whole genome’ SNPs. Geographically informative SNPs (iSNPs, *F*_ST_>0.05) show increased population clustering whilst randomly selected SNPs (rSNPs) and SNPs with similar allele frequencies (*F*_ST_<0.05) amongst different countries (universal, uSNPs) or local PNG populations (balanced, bSNPs) indicated little clustering. SNP panels must be validated in local settings to ensure they capture the diversity and population structure of the target population. Applied to 727 *P. falciparum* isolates from 16 provinces of PNG, the full barcode identified variable transmission dynamics, and eight major sub-populations, as well as the source of a malaria outbreak in a low transmission setting.

## MAIN

Pathogen genomics has become an essential tool for public health to detect, track and contain infectious disease threats ^1^. Whole genome sequencing (WGS) is a popular approach for small genomes such as those of viruses and some bacteria. However, for pathogens with larger genomes such as the malaria parasite, *Plasmodium falciparum* ^2^, targeted genotyping approaches are more affordable and scalable. Panels of single nucleotide polymorphisms (SNPs) referred as “SNP barcodes” have been increasingly used for malaria genomic surveillance ^3–8^. For malaria genomic surveillance to reach its full potential however, assessment of the capabilities and resolution of these genotyping tools is essential^9^.

Malaria remains a major public health problem for over half the world’s population, particularly for low- and middle-income countries in tropical and sub-tropical regions ^10^. Genomic surveillance of the causative Plasmodium parasites can play a critical role in malaria control and elimination, including for the quantification of local transmission dynamics, mapping population structure and connectivity, and identification of emerging or imported strains that may undermine control efforts ^6,11–20^. As malaria incidence declines, transmission becomes more heterogeneous, resulting in reduced gene flow between areas, and increasing focal inbreeding and relatedness of strains as the population size decreases ^16,21–24^. Maps of transmission zones, population connectivity and identification of “source and sink” populations could define the spatial scale and importation risk of distinct transmission zones and guide the containment of antimalarial drug resistance should it emerge ^25,26^. In pre-elimination areas, genotyping of parasites has been used to determine whether infections were imported or locally acquired, to characterise transmission chains and to guide outbreak preparedness and response ^27^. Genomic surveillance can therefore be used to monitor malaria control and elimination progress, and to improve the efficiency of control efforts ^20,28,29^.

It is crucial that genotyping tools used in malaria surveillance accurately define parasite population structure. Previous approaches have included panels of 10-14 polymorphic microsatellites ^11^, or barcodes comprising 24 ^30^, 28 ^3^, 54 ^31^ or 101 single nucleotide polymorphisms (SNPs)^6^. However, these genotyping tools lacked the required accuracy or spatial resolution to define local parasite populations, especially where the barcodes contained too few SNPs to accurately capture genome-wide relatedness ^17,32–34^. Indeed, the occurrence of low minor allele frequencies (MAF) of included SNPs in some populations reduces the spatial resolution of SNP barcodes. The development of a barcode that captures the diversity and divergence of local parasite populations may provide the most accurate and relevant insights for local malaria control programs ^35^.

We developed a SNP barcode to characterise parasite transmission dynamics throughout the Southwest Pacific country of Papua New Guinea (PNG). PNG is a major hotspot of malaria in the Asia Pacific, with heterogeneous *Plasmodium falciparum* transmission in different provinces ^36^. Control efforts intensified since 2006 reducing transmission and malaria burden in the period up to 2016 ^37–39^ with a national commitment to elimination from 2025 ^40^. We previously showed that *P. falciparum* populations were structured into subpopulations in 2005-6, prior to intensification of malaria control ^22,41,42^ which suggested that genomic surveillance could potentially identify transmission zones for targeted elimination. A barcode was designed using SNPs identified among *P. falciparum* isolates collected from three endemic areas of PNG. We then barcoded samples collected in a nationwide malaria indicator survey and from an outbreak of unknown origins in a low transmission area near the capital city, Port Moresby. The barcodes differentiated between local and imported infections and provided insights into the transmission dynamics, population structure and connectivity of *P. falciparum* parasite populations throughout PNG.

## RESULTS

### Barcode design and validation

SNP candidates were identified among 91 high quality *P. falciparum* whole genome sequences (WGS) from Milne Bay, East Sepik and Madang Provinces of PNG (**Figure 1A,B, Figure S1**, **Table S1)**^43,44^. This ‘local’ SNP barcode included 95 geographically informative SNPs with mean *F*_ST_ values among the three parasite populations of 0.05 or more considered to be geographically informative (iSNPs) and 97 SNPs with balanced allele frequencies based on *F*_ST_ values of less than 0.05 (bSNPs), and including 56 SNPs from an expanded version of a ‘Universal’ barcode that also met the above criteria (uSNPs) ^30, 31^ (**Figure S2, Table S2**).

**Figure 1.**
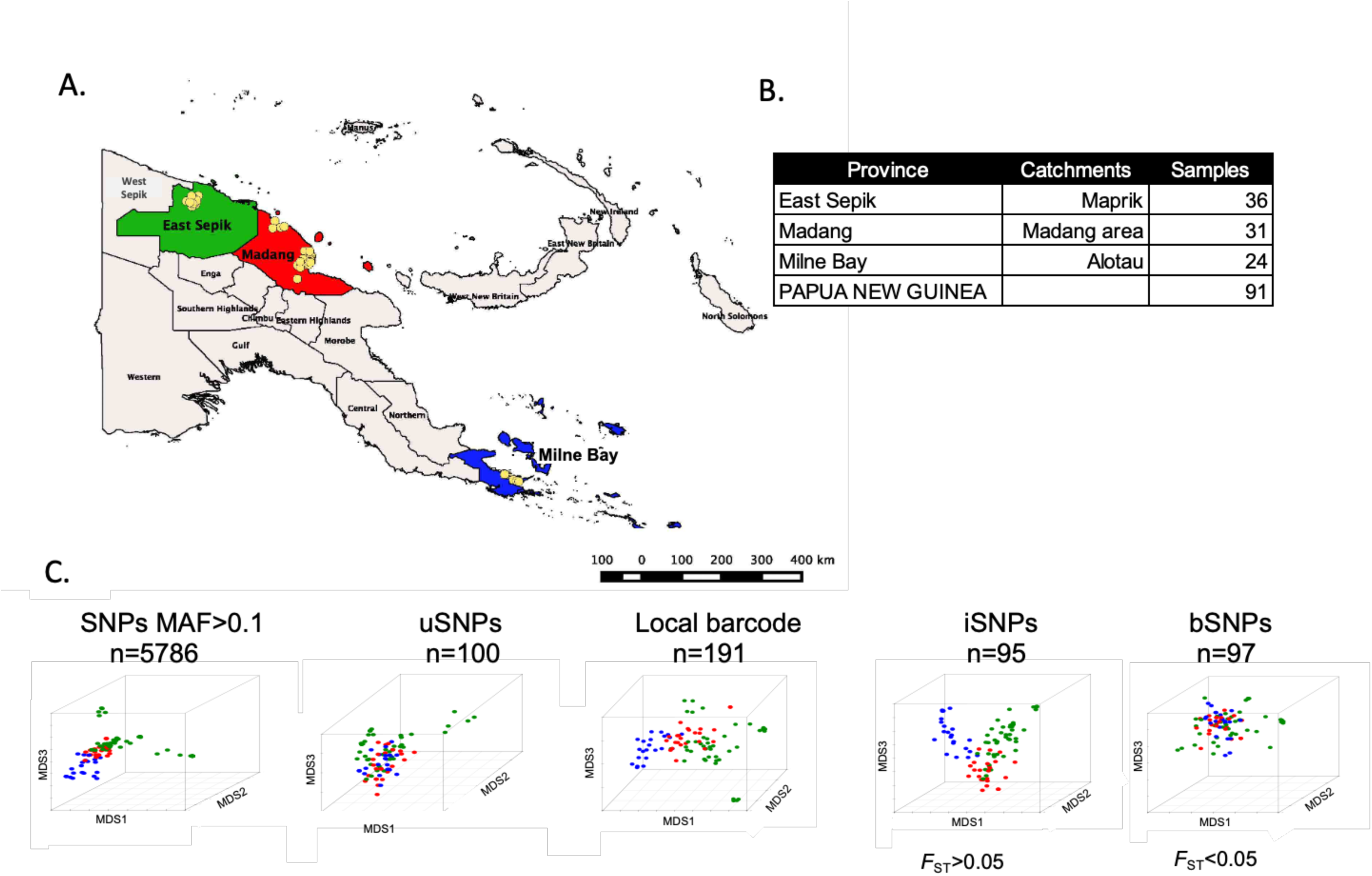
Clustering patterns of P. falciparum isolates from three PNG provinces based on different barcodes. A) Map of PNG indicating the village of residence of sample donors (yellow dots) and provinces (green = East Sepik Province, red = Madang Province, blue = Milne Bay Province). B) Number of high-quality P. falciparum genomes used for the analysis. C) Multidimensional Scaling (MDS) plots showing clustering patterns of different barcodes (n = number of SNPs). Coloured dots in the MDS correspond to the province in which samples were collected.

We compared barcodes comprised of all SNPs with minor allele frequency (MAF) of greater than 0.1 in PNG representing WGS (n=5786 SNPs), the uSNP barcode (n=96), the local barcode (n=191), iSNPs (n=95) and bSNPs (n=97) (**Figure 1C**). With all SNPs, Milne Bay isolates formed a distinct cluster while East Sepik and Madang isolates overlapped indicating significant gene flow between these neighbouring provinces (**Figure 1C**). Outliers observed in East Sepik cluster with isolates from neighbouring West Papua (data not shown, R. Amato, personal communication). The uSNPs and bSNPs failed to identify any population structure, while the iSNPs showed greater resolution of isolates by geographic origin. The improved resolution of the iSNPs suggests a trade-off between the number of SNPs in barcodes and their allele frequency characteristics (**Figure 1C**).

### Barcode performance

*P. falciparum* isolates from 68 villages across 16 of the 20 provinces of PNG (**Figure 3A,B**) were genotyped using the local barcode (**Table S2**). The cleaned dataset comprised 636 samples genotyped at 155 SNPs, including 73 iSNPs, 81 bSNPs, 46 uSNPs (**Table S2**). A random (rSNP) panel was also selected by combining 35 iSNPs and 35 bSNPs. The data was pooled with corresponding barcodes extracted from the WGS above providing 727 high quality SNP haplotypes in total. The dataset was stratified according to province (n=16) or catchment areas, assigned as ‘geocodes’ by combining proximal villages (n=34) (**Figure S3, Table S3**). The majority of samples within the final dataset had less than 10% of SNPs missing (**Figure S4**).

The population structure using different barcodes showed geographical structure only for the full barcode and iSNPs (**Figure 2C,D**). The rSNPs captured some population structure but did not identify Milne Bay as a distinct population, which was at odds with the ‘all SNPs’ data (**Figure 1**). The uSNPs and bSNPs showed the least population structure in PNG (**Figure 2C,D**).

**Figure 2.**
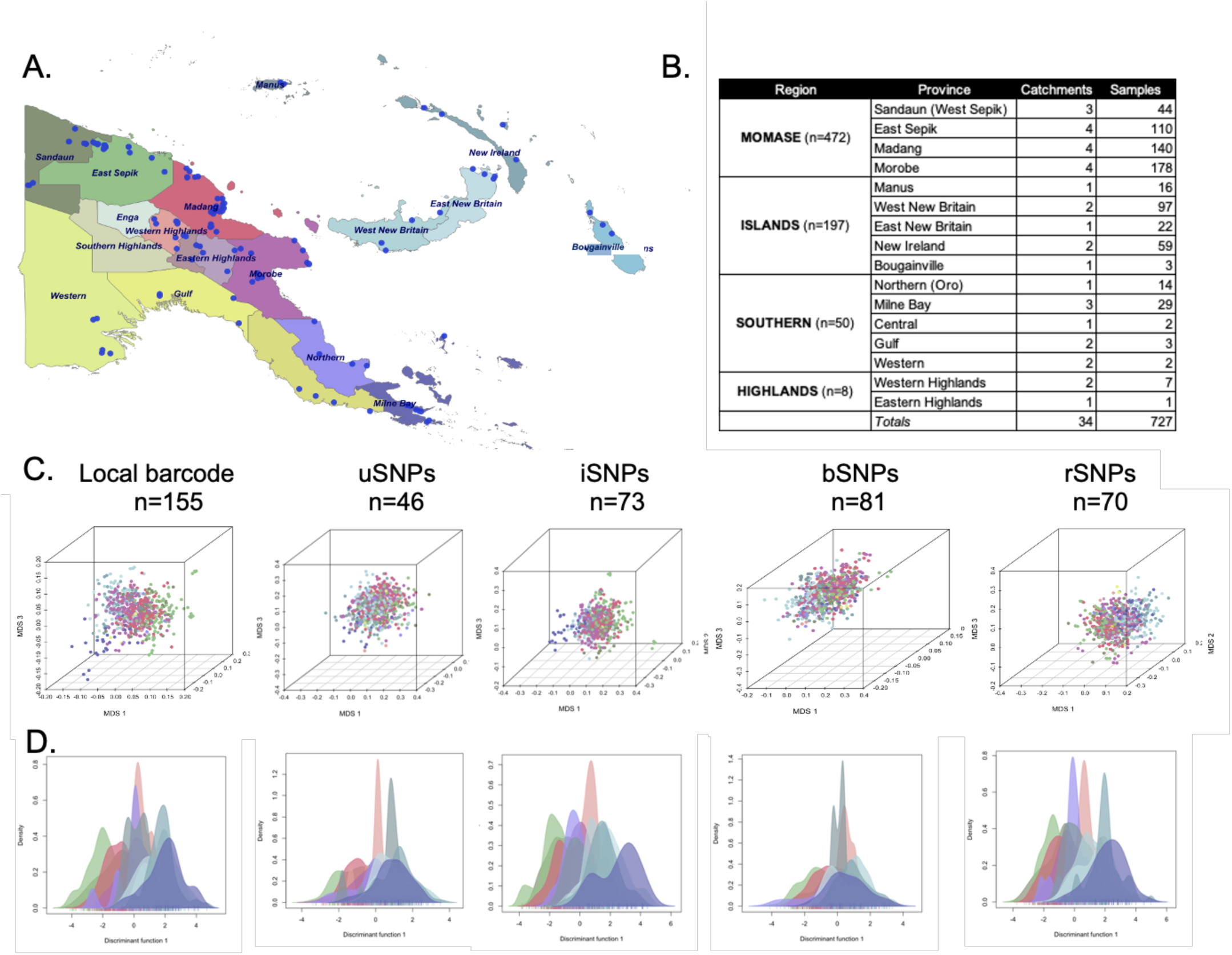
SNP barcode performance on Papua New Guinean P. falciparum isolates. A) Map of Papua New Guinea with sampling locations. Dots indicate villages, colours indicate different provinces. B) Numbers of catchment areas and successfully genotyped samples for each of the endemic provinces. C) Multidimensional Scaling (MDS) analysis of the samples using different panels of SNPs. n= number of SNPs. D) Discriminant Analysis of Principle Components (DAPC) density plots showing the dispersion of pairwise diversity values for populations with at least 7 samples. Colours correspond to provinces as indicated on the map.

### Variable transmission dynamics across PNG

Having confirmed the performance of the local barcode, we then determined the transmission dynamics of *P. falciparum* populations in PNG, using the complete panel of ‘typable’ SNPs (n=155). Population nucleotide diversity (ρε) and pairwise relatedness (expected identity by descent, eIBD) ^45^ was measured. Nucleotide diversity was high in all populations across the country, though slightly lower in the island populations suggesting lower transmission or a relatively isolated population (**Figure 3A**). Overall, PNG parasites showed low relatedness as expected for a high transmission, endemic, setting with frequent recombination. More related pairs were found in West Sepik (geocodes 1, 2), East Sepik (geocode. 4), Manus (geocode 19) and other island populations (geocodes 20-26) (**Figure 3A**). Larger proportions of closely related pairs (eIBD>0.55) were found in East Sepik (geocode 4) and Manus (geocode 19) suggesting clonal or near-clonal transmission (**Figure 3A**). Central Province (samples collected east of the capital city, Port Moresby), and Gulf Province had small sample sizes (n=2) due to the low prevalence of *P. falciparum* in these areas ^36^ and therefore were not shown on the plot. Central Province parasites were not related (eIBD<0.50) suggesting they were imported from distinct sources, whilst Gulf Province genotypes were identical, consistent with their originating from the same source and local transmission of this clone (data not shown in Figure 3A due to small sample size).

**Figure 3.**
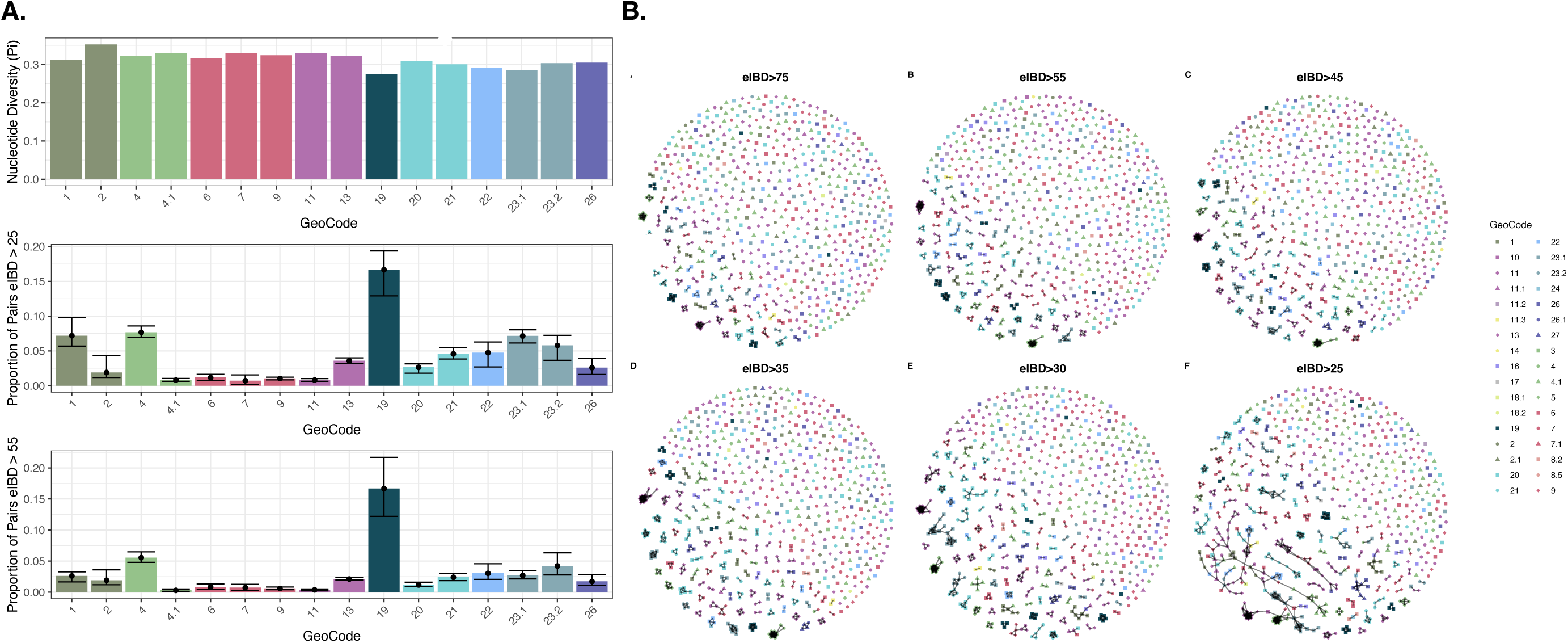
Diversity and relatedness of Papua New Guinean P. falciparum isolates. A. Within population nucleotide diversity (ν) (top) and relatedness (proportion of pairs with Expected Identity by Descent parameter (eIBD) > 25% (middle) and 55% (bottom). Error bars indicate 95% confidence intervals after bootstrapping (500 iterations). Only geocodes with 15 or more samples were included in this analysis. **B.** Networks of within population connectivity for different eIBD cut-offs. Nodes indicate parasite genotypes, coloured by geographical origins and lines indicate pairs with significant levels of IBD with a threshold of A) 0.75, B) 0.55, C) 0.45, D) 0.35, E) 0.30 and F) 0.25. Names of geographic catchment areas numbers in the key are shown in Table S4. Colours indicate province as shown in the map in Figure 2A. Geocode names (province: area) are: 1. West Sepik: Yapsie, 2. West Sepik: Aitape, 2.1 West Sepik: Nuku, 3 East Sepik: Yangroru, 4. East Sepik: Maprik, 4.1. East Sepik: Dreikikir, 5 East Sepik: Angoram, 6. Madang: Bogia, 7 and 7.1. Madang: Madang, 8.2 Western Highlands Province: Jimi, 8.5 Western Highlands Province: Wipim, 9. Madang: Sausi+Ramu valley, 10. Morobe: Markham valley, 11. Morobe: Mumeng, 11.1. Morobe: Bololu, 11.3. Gulf: Karema, 13. Morobe: Huon Peninsula, 14. Gulf: Kikori, 16. Oro/Northern, 17. Central: coastal, 18.1 Western: Balimo, 18.2 Western: Wipim, 19. Manus, 20. West New Britain: Kimbe Bay, 21. West New Britain: South Coast, 22. East New Britain: Gazelle Peninsula, 23.1. New Ireland: Namantanai, 23.2. New Island: Kavieng, 24. Bougainville, 26 and 26.1. Milne Bay: Alotau, 27. Milne Bay: Kiriwina. More details on the geocodes are available in Table S3.

Haplotype networks created using the pairwise eIBD values identified several clusters of highly related (eIBD>0.55) and closely related (eIBD>0.75) parasites (**Figure 3B**). Most clusters were comprised of parasites from the same geographic area, with only one cluster containing isolates from different geographic areas, suggesting importations. At eIBD thresholds above 0.75, clusters are detected within villages or between neighbouring villages only. As eIBD thresholds were relaxed from 0.55 to 0.45 to 0.35, relatedness remained within the villages, or between neighbouring villages only, indicating that lower eIBD thresholds are necessary to resolve population connectivity at sub-provincial spatial scales. At an eIBD threshold of 0.30, there was evidence of connectivity between neighbouring provinces within Momase and the Outlying Islands, and limited evidence of relatedness between more distant provinces. More extensive evidence of population connectivity was conspicuous at an eIBD threshold of 0.25, however this is the lower limit of resolution barcodes and prone to higher false-positive rates (unpublished data).

### Eight genetically distinct parasite sub-populations in Papua New Guinea

A spatial “gradient” of parasite population structure was observed for the north coast (West Sepik (Sandaun) > East Sepik > Madang > Morobe) with admixture between neighbouring provinces (West Sepik(Sandaun)/East Sepik, and Madang/Morobe, **Figure 4A**) accompanied by low pairwise *F*_ST_ values (0.01-0.05, **Figure S5**), consistent with a single contiguous parasite population with individual catchment areas isolated by distance. The outlying Islands (East and West New Britain, New Ireland, Manus Island) and Milne Bay isolates showed limited overlap and were genetically distinct to mainland isolates (mean *F*_ST_ = 0.03-0.18, **Figure S5**). STRUCTURE analysis identified eight genetically distinct clusters with three major regional blocks on the mainland comprising genotypes from East/West Sepik, Madang/Morobe and Milne Bay, distinct populations for Manus Island and the Islands (East and West New Britain and New Ireland) (**Figure 5B**) and multiple populations in Morobe, with Huon Peninsula isolates sharing ancestry with Island isolates (**Figure 5B**). There were some island genotypes with shared ancestry with the north coast (green and red) indicating recent importations from provinces on the north coast. Highlands and South Coast isolates shared ancestry within either Island, West and East Sepik or Madang/Morobe populations, consistent with multiple importation events.

**Figure 4.**
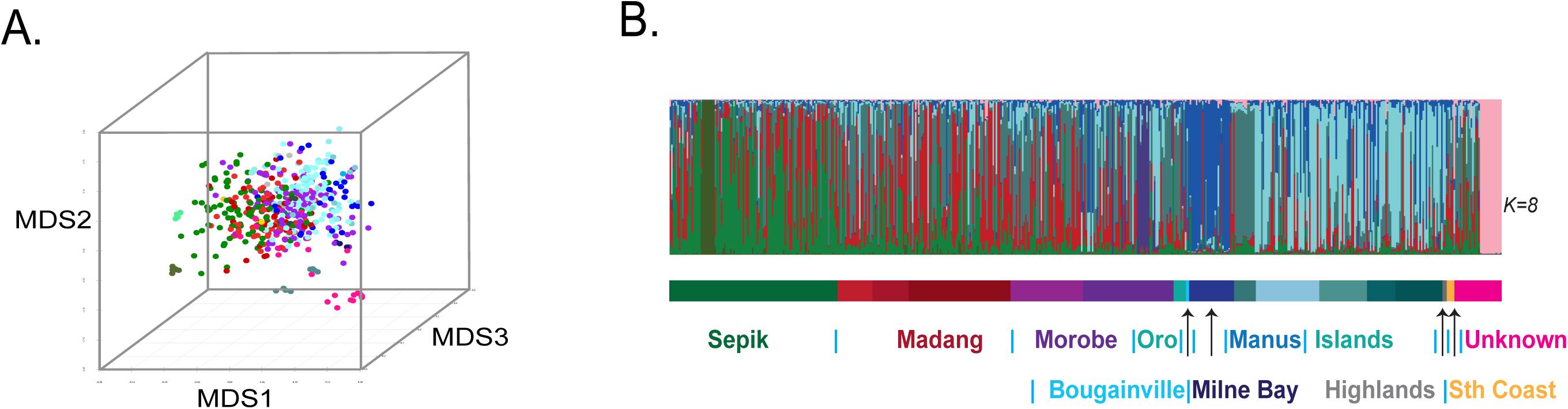
Population structure of P. falciparum in Papua New Guinea. A) MDS analysis of the outbreak samples with all PNG samples. B. Ancestry coefficients (min = 0 max =1) of outbreak samples with all PNG samples using Bayesian cluster analysis. Colours match geographical origins as indicated on the map in Figure 2A. The coloured bars below indicate collections of isolates from ‘Sepik’ which includes East Sepik and West Sepik (also known as Sandaun), ‘Oro’ is also known as Northern Province, ‘Islands’ includes isolates from East and West New Britain, and New Ireland, ‘Highlands’ combines all isolates from highland areas including Western Highlands and Eastern Highlands, ‘South Coast’ includes isolates from Central, Gulf and Western Provinces, unknown genotypes (pink) correspond to clinical samples of unknown geographical origin from migrating workers as described in the next section. Further details of these locations can be found in Table S3.

**Figure 5.**
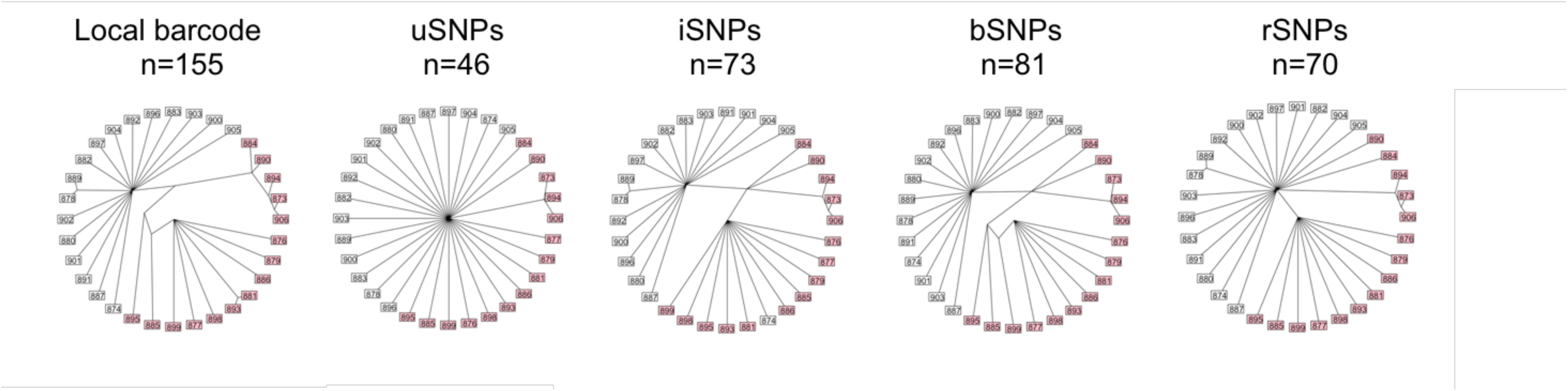
Phylogenetic analysis of P. falciparum haplotypes from a malaria outbreak using different SNP barcodes. Neighbour joining trees for different SNP barcodes. Neighbour joining trees for different SNP barcodes were created using the bionj function in R statistical software. Bootstrapping was performed by collapsing weak nodes with bootstrap support less than 70% over 1000 iterations. Samples indicated in pink were identified to be part of the same cluster by IBD analysis, using a threshold of eIBD>0.45 to connect samples (only one distinct cluster was detected) (**Figure S6**).

### Identifying the origins of infections in a malaria outbreak

To assess whether SNP barcodes with different characteristics can provide information on the source of outbreaks, we genotyped 32 *P. falciparum* clinical isolates from cases collected during a malaria outbreak that occurred amongst migrant workers over a 12-month period and compared them to the national dataset as a reference.

A subset of the outbreak samples were closely related outliers to the national dataset, whilst others clustered with samples from other endemic areas include in this study (**Figure 4A,B**). Detected eIBD connections for eight isolates showed low eIBD values of between 0.25-0.29 (**Table S5**). Three of the 15 isolates appeared to be related to isolates from the south coast of West New Britain and the Huon Peninsula of Morobe Province, suggesting that the outbreak was seeded by imported cases from these provinces. The remaining 17 haplotypes within the outbreak network were diverse and exhibited limited eIBD sharing with other infections within the outbreak.

The local barcode, iSNPs and bSNPs showed similar clustering patterns, dividing related (eIBD>0.45, Figure S6) infections (pink) into a distinct clade from the more diverse imported infections (**Figure 5**). However, uSNPs were only able to identify three related infections, and the rSNPs indicated two independent sources of the outbreak. The phylogenies identified a monophyletic clade with 15 related haplotypes suggesting a clonal expansion from genetically related parasites (Figure 6, pink nodes) with eIBD values of more than 0.45 (**Figure S6**).

**Figure 6.**
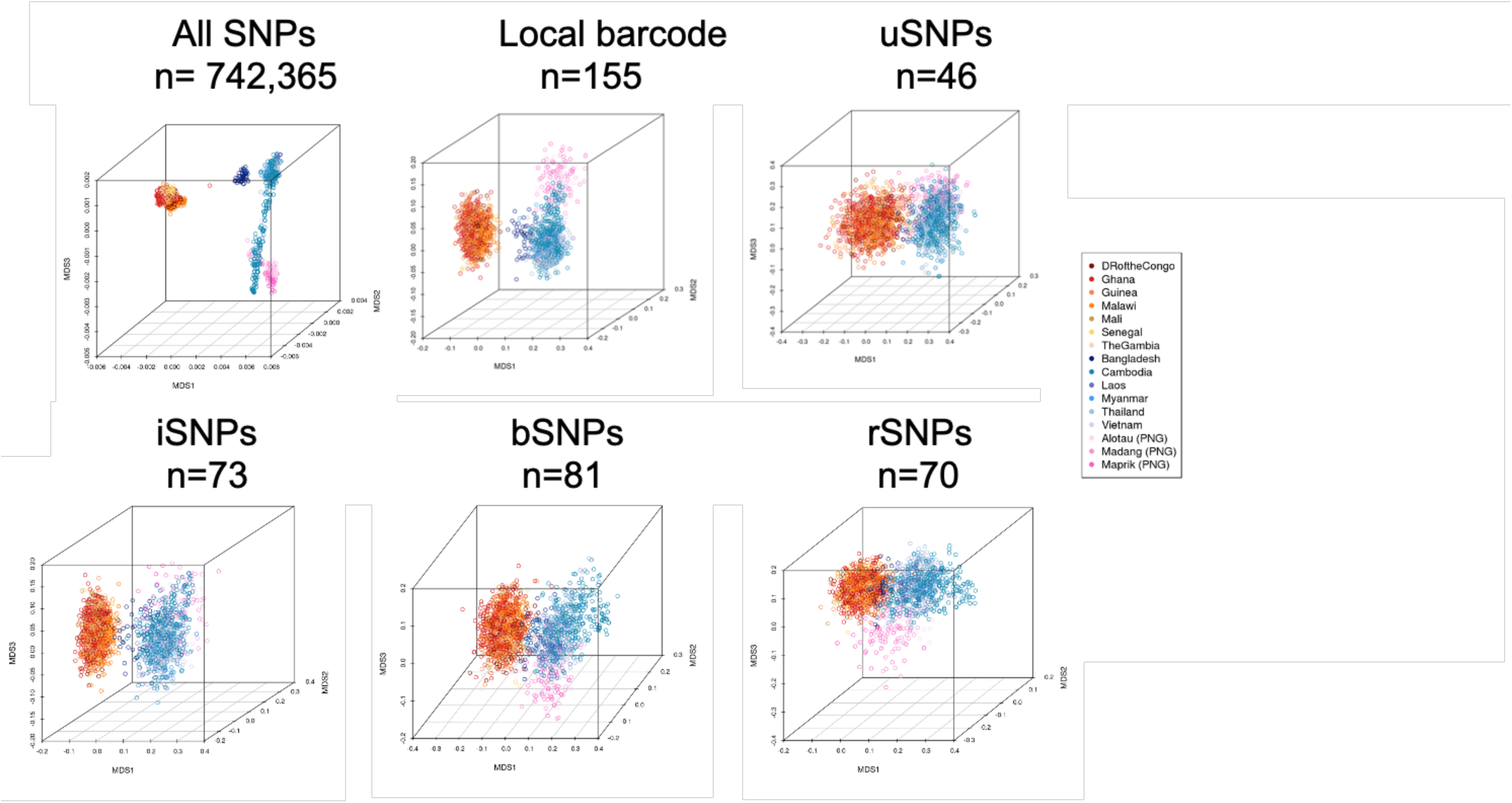
Clustering patterns of worldwide P. falciparum isolates based on different SNP barcodes. SNP genotypes from Pf3K v5.0 genomes were combined with data from the three PNG populations (n=2789 isolates). Isolates with genotype missingness <20% across the relevant panel of SNPs were selected, and MDS analysis conducted for all high-quality SNPs in the Pf3k dataset (n=742365 SNPs, n=2598 isolates), and validated SNPs from the local barcode (n=155 SNPs, n=2630 isolates), uSNPs (n=46 SNPs, n=2628 isolates), iSNPs (n=73 SNPs, n=2632 isolates), bSNPs (n=81 SNPs, n=2629 isolates) and rSNPs (n=70 SNPs, n=2628 isolates).

### Utility of the barcode for other malaria endemic regions

We also sought to assess the applicability of this SNP barcode to define parasite populations in other geographic regions. The 155 validated SNP loci were extracted from a global *P. falciparum* WGS dataset (Pf3k version 5.0 from 14 countries ^46^ in addition to PNG isolates described above [44,43] (Total n=2598 isolates). Based on 742,365 high-quality SNPs identified in this dataset, parasites clustered into broad continental regions, i.e. Africa, Southeast Asia, South Asia (Bangladesh), Oceania (PNG) (**Figure 6**). Most SNPs in the barcode were polymorphic in parasite populations of these regions, with at least 90 SNPs showing MAF>0.05 in African and Southeast Asian countries (**Figure S7, Table S6**).

The MDS showed similar population structures irrespective of the SNP panel used, albeit with type-specific clinal differentiation (**Figure 6**). Barcodes based on bSNPs and rSNPs differentiated PNG parasite populations from Asia Pacific and Africa, whilst uSNPs and iSNPs differentiated African and Southeast Asian isolates only, with PNG and Bangladesh showing clinal differentiation to Southeast Asian populations (**Figure 6**). This pattern of differentiation suggested that the iSNPs alone lacked sufficient discriminatory power for other Asia-Pacific regions.

## DISCUSSION

SNP barcoding can be done at a fraction of the cost of WGS and has utility for detecting and tracking malaria transmission dynamics in different endemic areas. Here we developed a SNP barcode tailored to capture the diversity of the local PNG *P. falciparum* population, and to assess the capabilities of SNP barcodes with different allele frequency characteristics. A SNP barcode comprised of 155 typable genome-wide SNPs was used to define the local parasite population structure and has utility to track outbreaks among clinical cases in PNG. Our results also suggest the SNP barcode could be used for different endemic regions globally, albeit refinements in the SNP panel may be required to ensure optimal results. Our SNP barcode together with the data generated in this study will inform control activities conducted by the PNG national malaria control programme.

A universal SNP barcode (uSNPs), designed using sequence diversity in non-target populations, may lack sufficient discriminatory power to define local parasite populations. Previous studies suggested that barcodes of 100-200 SNPs can provide accurate measures of genome wide IBD for measuring population connectivity ^34^. Our aim was to develop a barcode that would have broad practical and geographical utility, typical of SNP allele frequency characteristics within three ‘reference’ high transmission parasite populations of PNG. Geographically informative SNPs (iSNPs) were included to maximise classification of sample origins at sub-national scale in PNG (*F_ST_*>0.05, iSNPs), whilst others had balanced allele frequencies between the reference populations (*F_ST_* <0.05, bSNPs). SNP characteristics were representative of WGS, to provide an accurate estimate of underlying transmission dynamics and population structure using all typable local barcode SNPs, with a subset of SNPs for determining geographic origins of individual malaria cases (iSNPs). Selectively neutral SNPs were chosen to avoid drug or immune pressure that could skew observed patterns. Differences among SNP panels were dependent on SNP number, and allele frequencies within (MAF), and between (*F*_ST_), populations.

The iSNPs barcode identified increased population structure with fewer SNPs, however the randomly selected panel known as rSNPs, and the uSNPs/bSNPs with balanced allele frequencies, were not able to define the population structure observed using WGS. Therefore, refined barcodes as used in this study, and validation of SNP barcodes against WGS is a more cautious approach than the random selections of markers. The cost could be reduced by reducing the number of SNPs in a panel. However, this could also compromise the genetic resolution in parasite populations, with limited resolution in high compared to low transmission settings ^17,35^. In regions with higher transmission such as those in PNG and sub-Saharan Africa with little-to-no LD in the parasite population, accurate IBD estimates will require larger SNP panels ^35,47^ and should be assessed against WGS for ascertainment bias ^8^. In under-resourced settings, smaller panels of SNP barcodes could be tailored to the local parasite populations and the research question. Given the limited access to high-throughput genotyping platforms in endemic countries, optimised SNP barcodes could be operationally feasible for specific malaria control use cases.

The validated local barcode was suitable for characterising subnational population structure and transmission dynamics and showed utility to track local outbreaks and imported cases. As only low levels of relatedness were detected between geographic areas, levels of connectivity were not inferred using eIBD methods as others have done ^16,32^. Instead, we base the inference of connectivity between populations on *F_ST_* values and levels of admixture or shared ancestry in the STRUCTURE analysis. Three major population subdivisions were found within PNG: the mainland-north coast (known as the Momase region), Eastern Tip (Milne Bay Province) and outlying Islands, in addition to distinct populations for Manus Island, Northern (Oro) and Central Provinces, and highly focal sub-populations on the Morobe coast. Previously, parasite population structure was observed on the north coast between East Sepik and Madang using microsatellite markers ^15,22,41^. This geographic population structure may be explained by limited human movement at the time of sample collection and may indicate distinct malaria transmission ‘zones’, which defines the spatial scale that may be needed for targeted control efforts.

Morobe parasites were structured on a fine scale, according to village origins with some evidence of importation from the Islands to the Huon Peninsula which received regular boat traffic from the islands. This indicates a potential transmission zone boundary and migration point between Islands and the Mainland, that should be considered in subnational control efforts. Evidence of imported infections were also observed in the East Sepik with a cluster of isolates that were outliers to the entire PNG parasite population. Recent studies have indicated significant mixing among parasite populations in Wewak, East Sepik and from the West Papuan (Indonesian) side of the island of New Guinea ^48^. Microsatellite data from East Sepik indicated focal transmission with clusters of related parasites within certain villages^22^. East Sepik implemented multiple LLIN distributions prior to this study, suggesting focal clustering suggesting interrupted transmission relative to other north coast provinces, and these imported infections may pose an additional risk to malaria elimination.

Sample sizes from the Southern and Highlands regions were very small, limiting the conclusions made about these parasite populations. Relatedness estimates however suggested importation (not related) and/or clonal transmission (related). Low nucleotide diversity and high relatedness between genotypes from the Gulf Province suggests clonal transmission within the local area. Conversely, in Western, Central, and the highlands provinces, low parasite relatedness and similarity to parasites from highly endemic areas, suggests importation may be a major contributor to local malaria episodes. Low transmission areas like the Highlands and Southern regions may be potential sinks for infections, with the major populations in higher transmission areas of Momase and Islands are likely to be the major sources.

While it is important to maintain surveillance locally in each endemic province, there is also a need to monitor for imported cases that may potentially seed transmission in provinces moving towards elimination. The SNP barcode provided utility to detect and track the origins of local outbreaks in low transmission settings. Barcoding of the clinical isolates obtained from migrant workers revealed a local outbreak that spread from two origins: a single clade with an ancestor that was likely imported and transmitted to other individuals, and multiple imported cases. Our data suggested that the outbreak was seeded by two independent transmission events with a cluster of parasites that were likely imported from the north coast and island populations.

SNP candidates were identified from three parasite populations including one relatively geographically isolated population (Milne Bay). It is possible that the selection of SNPs based on mean *F*_ST_ among these three populations may have identified SNPs that are private to one of these populations, however none of the SNPs chosen were private to any population (**Table S2**). Genotyping success can be a limitation in barcoding studies, with the loss of almost 25% of the candidate SNPs. Some markers were removed due to batch effects, and others because they had a less than 70% genotyping success rate amongst the samples. This could be improved by validating additional SNP candidates, however mathematical modelling has indicated that 100-200 SNPs are adequate for IBD estimation ^34^ and our results clearly show the utility of the 155 remaining SNPs for describing the population genetics of *P. falciparum* in PNG, and possibly in other countries. PNG samples used were archival and may have reduced DNA quality with multiple freeze thaw cycles. They were also mostly from asymptomatic infections with low densities contributing to genotype failures. Sampling bias towards the high transmission regions and very few samples from the relatively low transmission Highlands and Southern regions of the country were unavoidable. Sampling in these areas may be boosted through inclusion of cases from malaria clinics and hospitals, to confirm the origins and dynamics of transmission.

The performance of the different SNP barcodes in PNG and among other malaria endemic countries within and outside the Asia-Pacific underscores the need to optimise SNP panels for local and/or regional parasite populations, factoring in the locus-specific MAF and *F*_ST_ estimates, to ensure subnational parasite population structure can be detected and monitored accurately. Integration of the spatial information provided by SNP barcodes into disease surveillance maps will be useful to define regions for targeted elimination, to measure the success of control interventions, detect and track the spread of drug resistant parasites and enhance outbreak preparedness across different transmission settings. Considering the recent recognition that new tools will be required to progress beyond the current stagnating progress against malaria, this research provides critical new evidence supporting the use of parasite genomic surveillance using SNP barcodes.

## METHODS

### Ethics and Informed Consent

All samples were collected as part of ongoing studies in PNG investigating the impact of intensified malaria control efforts since 2004. All samples were collected with written informed consent from individuals or if children, consent was obtained from their parents, and guardians. Ethical approval for the study was obtained from the Papua New Guinea Institute of Medical Research Institutional Review Board (IRB 11/21, 12/29), the PNG Medical Research Advisory Council (MRAC 12/03, 13/08), the Walter and Eliza Hall Institute Human Research Ethics Committee (HREC 12/06, 13/14) and Deakin Human Research Ethics Committee (2020-282, 2020-283).

### Study design

The objective of this study was to assess population structure at genome-wide resolution using WGS, and a SNP barcode tailored to local PNG parasites. For WGS, we collected whole blood samples from clinical malaria cases, thus the sample size was limited by the clinical cases and samples available from the original study. For SNP typing we utilised *P. falciparum* qPCR-positive blood samples collected from households in 68 villages as part of a national malaria indicator survey, and symptomatic, laboratory-confirmed clinical cases from four sentinel sites, and during a malaria outbreak amongst migrating workers residing in a camp in a low endemic area. For the samples from the national survey, we aimed to produce dense spatial sampling in high endemicity areas with a maximum of 30 samples per village, and in low endemicity areas we genotyped as many isolates as were available. Data collection was stopped when this number of samples was reached for each village. Data was included only if fewer than 30% of allele calls were missing from a genotype (sample) or marker (SNP). Using Principal Components Analysis (PCA), ‘outlier’ SNPs and samples were identified, but only outlier SNPs were excluded, since ‘outlier’ samples were likely to be imported infections, and thus included in the investigation. The units of study were groups of samples (parasite populations) stratified at large to small spatial scales including (i) region (n=4, Momase, Islands, Highlands, Southern), (ii) province (n=16) and (iii) catchments that were defined as ‘geocodes’ (n=34) comprising samples from proximal villages.

### Samples and study area

PNG is located on the eastern half of the island of New Guinea in the WHO Western Pacific region just to the north of Australia and shares a border to the West with Indonesia (West Papua). It is a major hotspot for malaria in the Asia-Pacific region with intense year-round malaria transmission ranging from hyper- to holo-endemic in the lowland and coastal areas of the Momase and Islands and hypo-endemic to epidemic in the Highlands and the less populated or peri-urban Southern regions ^36^.

For whole genome sequencing (WGS), *P. falciparum* isolates were collected from individuals diagnosed with clinical malaria during (i) a severe disease case control study conducted in 2005 in the Madang area, Madang Province ^49^, and (ii) an antimalarial efficacy trial conducted in 2012-13 in Maprik, East Sepik Province (7 villages) and Alotau, Milne Bay Province (9 Villages)^50^. The Madang samples (n=55, from 3 villages) were sequenced in previous studies^51^. For Maprik and Alotau, new isolates were collected for this study, and have been described in a previous publication^50^. In summary, a total of 1 mL of fresh venous blood was processed using a CF11 filtration procedure to obtain purified erythrocytes for DNA extraction according to published protocols ^52^. More than 202 *P. falciparum* positive clinical samples from Alotau and Maprik were submitted for sequencing, of those, 157 met the quality control threshold, and 122 met the quality thresholds for inclusion in the study (Supporting Information: **Text S1**, ^43^). This included removal of one individual of the pairs the samples identified as clonal or closely related to another through multidimensional scaling (MDS) and known location of collection and familial relationship (siblings from the same household). SNP candidates were initially selected using 56 of the highest quality genomes (Maprik = 16, Madang = 22, Alotau = 18) and further validated using a total of 91 isolates as more data became available (Maprik = 36, Madang = 31, Alotau = 24, **Figure 1B, Table S1**).

*P. falciparum* isolates for the barcoding were selected from a nationwide malaria indicator survey conducted by the PNG Institute of Medical Research between October 2008 and August 2009 ^36^. The survey included the collection of 6646 finger prick dried blood spots from individuals of all ages through a household survey in 49 villages, in addition to 2290 samples from individuals residing in 19 villages across six sentinel sites (^36,37^, **Table S1, Table S3**). In total the sample set included 8936 samples from individuals living in 68 villages in 16 of the 20 provinces of PNG. For molecular diagnosis of *P. falciparum*, DNA was extracted from dried blood spots using Qiagen or Favorgen extraction kits and a semi-quantitative post-polymerase chain reaction, ligase detection reaction/fluorescent microsphere assay conducted as described ^53^. *P. falciparum* isolates identified were previously genotyped to determine multiplicity of infection (MOI) ^54^ and selected for SNP genotyping if only one clone was present (MOI=1). If sample number was low for a particular location, low complexity multiple clone infections were included (MOI=2). In the previous study, 3784 *P. falciparum* positive samples were identified by qPCR and of those, 2400 were successfully genotyped using the highly polymorphic marker *Pfmsp2* ^54^. We selected 772 *P. falciparum* isolates for genotyping (MOI1= 585, 75.7%; MOI2=187, 24.3%) aiming to maximise sampling density across different provinces. Data cleaning included removing samples with low SNP genotyping success rates (n=167 samples with more than 30% missing SNP loci) and removing SNP loci with low quality output and batch effects (n=34) from all samples. This resulted in 636 samples successfully genotyped at 155 SNPs. For the population-level analyses we combined villages at two different spatial scales: province (n=16) and geographic catchment areas known as geocodes (n=34) defined based on the spatial proximity of villages and local knowledge of transport networks and topographical features that may influence parasite population movement (**Table S3, Figure S3**).

To investigate infections of unknown origins, we barcoded 32 *P. falciparum* isolates from individuals diagnosed with clinical malaria based at a resource company camp approximately 20 km from the capital city, Port Moresby. Although malaria is at very low prevalence ^36^, this region is receptive to malaria transmission, and therefore the company has strict malaria control procedures in place. However, workers leave the camp for social activities and to return to their home province, where they may acquire malaria infections. Travel history was not available for the participants.

Positive control samples for the SNP genotyping included pure *P. falciparum* laboratory strains 3D7 and D10. Mixtures combining different ratios of these clones were made as standards to calibrate the Fluidigm raw data analysis and to enable the detection of major alleles in the case of multiple infections.

### SNP candidate selection and validation

#### Whole genome sequencing

Leucocyte-depleted clinical *P. falciparum* isolates were sent to the Sanger Institute (Hinxton, UK) for Illumina-based whole genome sequencing supported by the Malaria Genomic Epidemiology Network (MalariaGEN) *P. falciparum* Community Project ^51^. This data has now been made publicly available ^44^. Illumina short read data was processed to extract high quality variant calls by the MalariaGEN team ^51^. Samples with poor coverage or diagnosed as containing multiple infections as determined by *Pfmsp2* genotyping ^55^ and/or *F*_ws_ values of less than 0.80 ^56^ were removed. The missingness threshold was determined using Poisson regression model using Mallows or Huber type robust estimators with the missingness counts as the response variable, and a cutoff of greater than 70% missing genotypes).

#### SNP candidates

Variant calls from WGS of 137 PNG *P. falciparum* isolates ^44,57^(**Figure S1, Table S1**) were filtered to obtain 682,796 high quality SNPs based on the MalariaGEN *P. falciparum* Community Project, Release 3.1 ^51^. Whole genome SNP genotypes were filtered to remove all monomorphic sites amonst the PNG genomes, leaving 24,633 SNPs. SNPs with minor allele frequencies (MAF) greater than 0.1, calculated directly in R using the adegenet version 1.3-8 software ^58,59^ were selected for further assessment. Quality control was done by removing samples with high missingness or evidence of polyclonal infection with *F*_WS_ values of less than 0.80, ^56^, and removing SNPs in blacklisted regions which include highly variable telomeric regions and those containing polymorphic multigene families which prevents accurate mapping of reads ^60^. The final dataset included 91 WGS ‘genotypes’ each with 5786 SNP calls (**Figure 1**). A total of 2459 SNPs (42.5%) were found to be geographically informative (*F*_ST_>0.05) in PNG as determined by calculating mean pairwise *F*_ST_ for each SNP using Weir and Cockerhams F_ST_ in the R DiveRsity package ^61^ (https://github.com/kkeenan02/diveRsity) as well as GenePop version 1.2 ^62^. Geographic classification potential was further determined by logistic regression in R Statistical Software by identifying SNP loci with classification probabilities ^58^. To select candidates for development of a barcode, filtering included removing SNPs in regions of the genome with evidence of selection identified using Bayescan ^63^ and Tajima’s D test with VCF tools version 0.1.13 ^64^ and using a cutoff of two standard deviations from the mean in a sliding window analysis. We also avoided regions containing known antigens and drug resistance genes likely to be under selection pressure. To ensure candidate SNPs were unlinked, Chi Squared tests of independence were conducted retaining only the 30 most significant variables. SNPs were then split by chromosome and ranked by *F*_ST_ which was previously measured as described above. Half of the SNPs were then selected to have *F*_ST_ values of at least 0.05, which indicates moderate genetic differentiation with confirmed high classification probability for geographic origin using logistic regression (iSNPs) and the other half had *F*_ST_ values less than 0.05 with balanced allele frequencies between populations (bSNPs). We also screened all SNPs against an extended universal barcode panel of 171 universal SNPs (uSNPs) previously developed by N. Baro and S. Volkman (unpublished data, Harvard T.H. Chan School of Public Health) developed based on African and Asian parasite populations ^30^ that were also found to be polymorphic in the PNG population. Of these, 111 overlapped with the filtered WGS SNPs, 107 were polymorphic in PNG and 96 had a MAF greater than or equal to 0.1. Before selecting our final panel, we confirmed they were “typable” based on having at least 5X read coverage in the WGS assemblies.

The final panel of candidate SNPs were selected to represent both the distribution of geographically informative (iSNPs, *F*_ST_>0.05) and balanced allele frequencies (bSNPs, *F*_ST_<0.05), and to include as many uSNPs as possible with a relatively uniform distribution and no less than 10 kB apart. Based on these criteria, a total of 324 SNP candidates were submitted to Fluidigm for assay development, including 165 iSNPS and 159 bSNPs of which 68 were uSNPs. Of these, 279 passed the stringent primer selection process and 191 were then selected for assay development including 95 iSNPs and 97 bSNPs of which 56 were uSNPs. The panel of 192 SNP assays in total included one duplicate SNP. Of the 191 SNPs, 88 were nonsynonymous and 105 were synonymous, and each chromosome was represented by 8 to 14 SNPs (**Figure S2, Table S2**).

### SNP barcoding

SNP genotyping assays were developed for the final panel of 191 SNPs using the Fluidigm SNPType® system using the Fluidigm BioMark platform and 96.96 Dynamic Array Integrated Fluidic Circuit (IFC) genotyping (Supporting Information: **Figure S2B**), following the manufacturer’s instructions with minor modifications. Two IFC chips per batch of 84 samples were needed to genotype all SNPs in the barcode. Samples on each chip included a total of 84 field isolates, plus 12 controls including pure 3D7 and D10 DNA and mixtures in ratios of 90:10, 80:20, 70:30, 60:40, 50:50, 40:60, 30:70, 20:80 and 10:90, and a no DNA (water) template as the negative control. The assay was first optimised using the controls, and then used to genotype field samples.

#### SNP barcoding assays

Due to the low parasitemia of many samples and low volume required, 30 µL of extracted DNA was first reduced to 3 µL using a desiccator and 1.25 µL was added to each primary multiplex reaction containing all 191 primer pairs. Primary reactions were carried out in a total volume of 5 µL in 96 well plates on a T100, BIO-RAD thermal cycler using the following conditions: 2.5 µL (1x) Qiagen 2X Multiplex PCR Master Mix (Cat No:206143, USA) and 0.5uL (0.2 µM) each of the Specific Target Amplification (STA) & Locus Specific (LSP) primers. The cycling protocol included an initial denaturation for 15 min at 95 °C, followed by 16 cycles of 15 sec denaturation at 95 °C and annealing & extension together for 4 min at 60 °C. The secondary reaction is a qPCR end point fluorescence assay using Biotium 2x Fast Probe master mix (Cat No. 31005, Biotium, US), Rox (Cat No.12223-012, Invitrogen, US) and SNPtype genotyping reagent kit 96.96 (Cat No.100-4134, Fluidigm, US). Priming and loading of IFC into BioMark Platform was performed according to manufacturer’s instructions. IFC chips (Cat No. BMK-M-96.96GT, Fluidigm, USA) contain 96 separate reservoirs on each side of the microfluidic device, one for samples, into which 5uL of the sample mix containing 1:25 diluted primary PCR product and the other side 4 µL of assay mix containing Allele Specific Probes(ASP) and Locus Specific (LSP) primers were aliquoted following the manufacturer’s instructions (**Table S4**). Within the IFC, 9216 individual qPCR reactions are assembled automatically via microfluidics using the IFC controller. The cycling protocol used on the Biomark TM instrument was “SNPtype 96.96 v1(Fluidigm, US). Each IFC chip was set up to genotype 96 SNPs in 96 samples, which included different ration of mock multiple infection controls comprised of single and mixed proportions of the reference strains 3D7 and D10 (90:10, 70:30, 60:40, 50:50, 30:70, 40:60, 10:90).

#### SNP calling

Allele specific PCR products were generated and imaged with scatter plots using Fluidigm SNP genotyping analysis Software (Fluidigm, United States). Briefly, a scatterplot is produced for all samples on the chip per SNP to enable automated allele calling by the software. In the case of multiple infections, the dominant allele was identified by the spectra/location on the scatter plot relative to dominant alleles in the control mixed infections (**Figure S2B**).

### Data analysis and statistics

SNP barcode data was analysed using multiple approaches. Briefly, nucleotide diversity (ρε) ^65^, was calculated using the function *nuc.div* in Pegas (V0.12) ^66^ with pairwise deletion of missing loci. Multilocus *F_ST_* was calculated for each pair of populations using the function *varcomp.glob* in Hierfstat (V0.04-22)_67._

Bayesian cluster analysis was done using STRUCTURE version 2.3 ^68^ using the following parameters: K=1 to 20 with 20 runs with different seeds, an MCMC burn-in period of 5000 iterations and total MCMC iterations up to 50,000. Optimal K was determined by using the inflection point in final estimated log-likelihood and the ‘Evanno et al’ method ^69^. MCMC diagnostics were also checked by looking at change in admixture parameter over MCMC runs. Samples were labelled by population, but population origin was not set as a prior in the analysis to avoid overestimation of population structure.

Clustering and phylogenetic analyses for SNP data were based on genotypic distances. The distance between a pair of isolates was defined to be the proportion of loci with differing genotypes with pairwise deletion of missing loci, as implemented in the R package ape (V5.0) ^70^. Classical multidimensional scaling analysis (MDS), as described by ^71^, was performed on the pairwise distance matrix using cmdscale function in the base statistics package (V3.4.0) in R. Discriminant Analysis of Principal Components (DAPC) was also performed on the pairwise distance matrix using Adegenet (V2.1.1) ^59^, with isolates stratified into provincial populations. The number of principal components retained for DAPC was informed by the ‘a-score’ (defined to be the difference between observed and random discrimination) to optimise discrimination power and avoid excessive overfitting. Phylogenetic analysis of isolates collected during an outbreak in migrant workers was done by constructing Neighbour Joining trees using the BIONJ algorithm implemented in ape (V 5.0) ^70^. Bootstrapping was performed to collapse nodes with bootstrapping support below 70% over 1000 replicates.

Pairwise relatedness (IBD sharing) between isolates was inferred using *IsoRelate* ^45^, setting the genetic distance to be 1cM=13.5kbp ^60^ and assuming a genotyping error rate of 0.001. Briefly, the posterior probability of IBD sharing at each locus was averaged for each pair of isolates to obtain the expected fraction IBD (eIBD), ^32^; posterior probability estimates output by *IsoRelate* were corrected by a factor of 2. Adjacency matrices for various eIBD thresholds (namely, 0.25, 0.30, 0.35, 0.45, 0.55 and 0.75) were used to construct IBD networks using the R package *igraph* (V1.2.4.1) ^72^. As a measure of diversity, we calculated the proportions of pairs with eIBD sharing above 0.25 and 0.55 for each subpopulation. Bootstrapping (500 iterations) was performed over eIBD values to generate 95% confidence intervals for the proportion of pairs IBD.

## Data Availability

All raw whole genome sequence data is available at the European Nucleotide Archive using the accession numbers indicated in Table S1. The final SNP barcode dataset for 727 isolates and for 32 outbreak samples will be made available in a public repository upon publication, and in the interim will be available by contacting the corresponding author.

## Acknowledgements

We are grateful to the communities that participated in the study and the staff of the Papua New Guinea Institute of Medical Research that contributed to the field studies and sample collections. Dr. D. Hill and Dr. M. Hapsari Hazarin contributed technical assistance during the study and Drs. S. Volkman and N. Baro shared co-ordinates of uSNPs. Dr. M. Manske assisted with genomic sequencing logistics. Drs. K. McCann and C. Narh assisted with editing the manuscript. Sample collections, DNA extractions and molecular diagnosis were funded by the Global Fund to Fight AIDS Tuberculosis and Malaria. Genetic and genomic data collection for this was funded through a National Health and Medical Research Council (NHMRC) of Australia Project Grant Number GNT1027108. IM and MB are supported by NHMRC Research Fellowships (GNT1155075, GNT1102971). The authors acknowledge the Victorian State Government Operational Infrastructure Support and Australian Government NHMRC Independent Research Institute Infrastructure Support Scheme (IRIISS).

## Author Contributions

Conceived and designed the analysis: GLAH, SM, MB, IM, AEB; Collected the data; GLAH, SM, ZR; Contributed resources: MWH, LT, ML, LM, NB, AJ, IB, PMS; Contributed data or analysis tools: RA, OM, DK, MB; Performed the analysis; GLAH, SM, NT; Wrote the paper: AEB.

## Materials and Correspondence

Please address correspondence to Professor Alyssa Barry.

