## Supporting Information for "*Plasmodium falciparum* populations, transmission dynamics and infection origins across Papua New Guinea"

†deceased 27 April 2023

**SUPPORTING FIGURES**

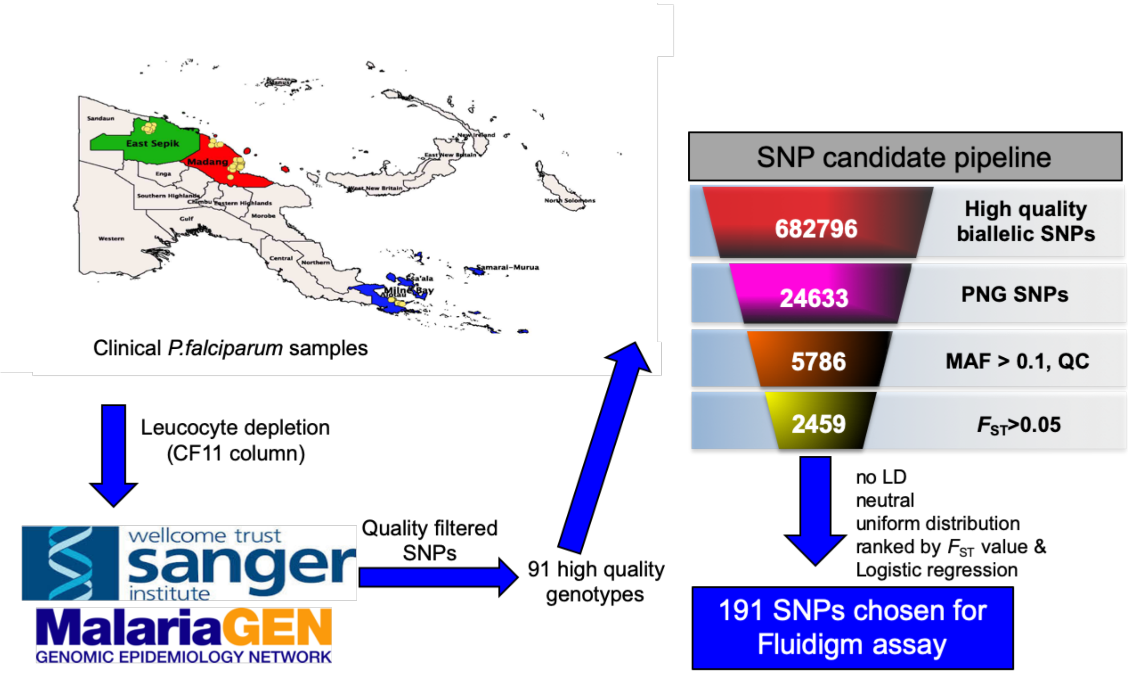


**Figure S1. Pipeline for selection of candidate SNPs.**

Whole blood samples from three locations of PNG (yellow dots) were subject to leucocyte depletion by CF11 filtration. DNA was extracted and sent to the Sanger Institute for sequencing as part of the MalariaGEN *P. falciparum* Community Project [1]. High quality biallelic SNP genotypes were returned comprising allele calls for 682,796 SNP loci. These were then filtered for polymorphic loci amongst the PNG samples with minor allele frequencies (MAF) greater than 0.1. A total of 42.5% (2459/5786) were classified as geographically informative (mean $F_{ST}$>0.05). Neutral, unlinked (no linkage disequilibrium, i.e., LD), relatively uniformly distributed SNPs were then selected with approximately 50% selected for geographic classification (logistic regression and/or $F_{ST}$).

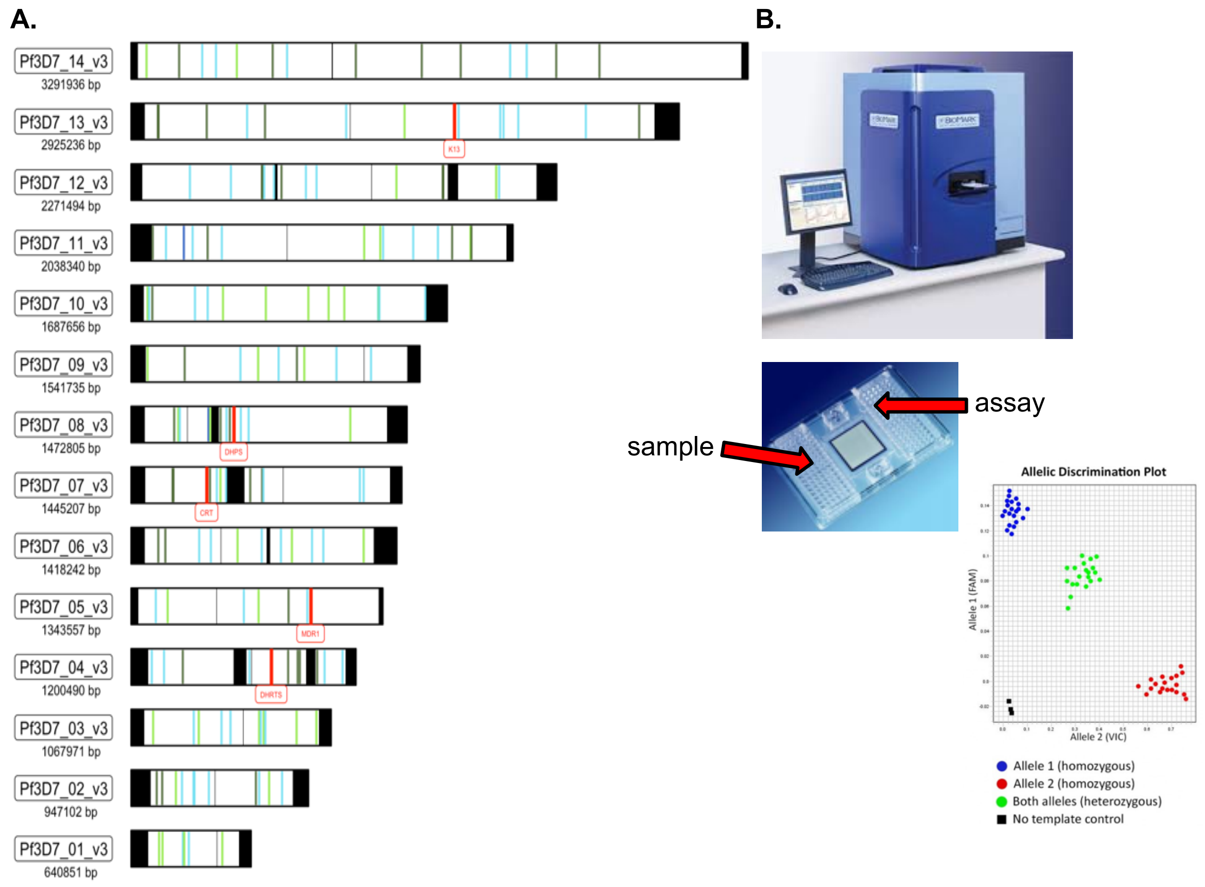


**Figure S2. SNP Barcode and Assay**

**A)** Chromosomal locations of barcode SNPs. Each chromosome is shown as a horizontal bar, with SNPs highlighted by blue ($F_{ST}$>0.05) and green ($F_{ST}$<0.05) bars. Darker colours indicate those positions that are also part of the universal barcode (uSNPs). Black bars indicate excluded regions. Red bars indicate locations of drug resistance landmarks (chr 4: *dhfr-ts*, chr5: *mdr1*, chr7: *crt*, chr8: *dhps*, chr13: *kelch13*). **B)** Fluidigm SNPType® Assay. Assays were done using the BioMark platform. Fluidigm IFC's contain 96 wells for samples, and 96 wells for assay master mix containing primers and two probes for each allele. The results are output as fluorescent scatterplots with dots representing samples with the reference allele shown in blue, and alternative alleles shown in red. Green dots indicate samples that have a mixture of both alleles (heterozygous). Each experiment (IFC chip) included pure DNA and mixtures of varying ratios for *Plasmodium falciparum* 3D7 and 7g8 clones and therefore plotted as a spectrum between samples with reference and alternative alleles. Only samples with clear dominant allele calls were included.

**A.**

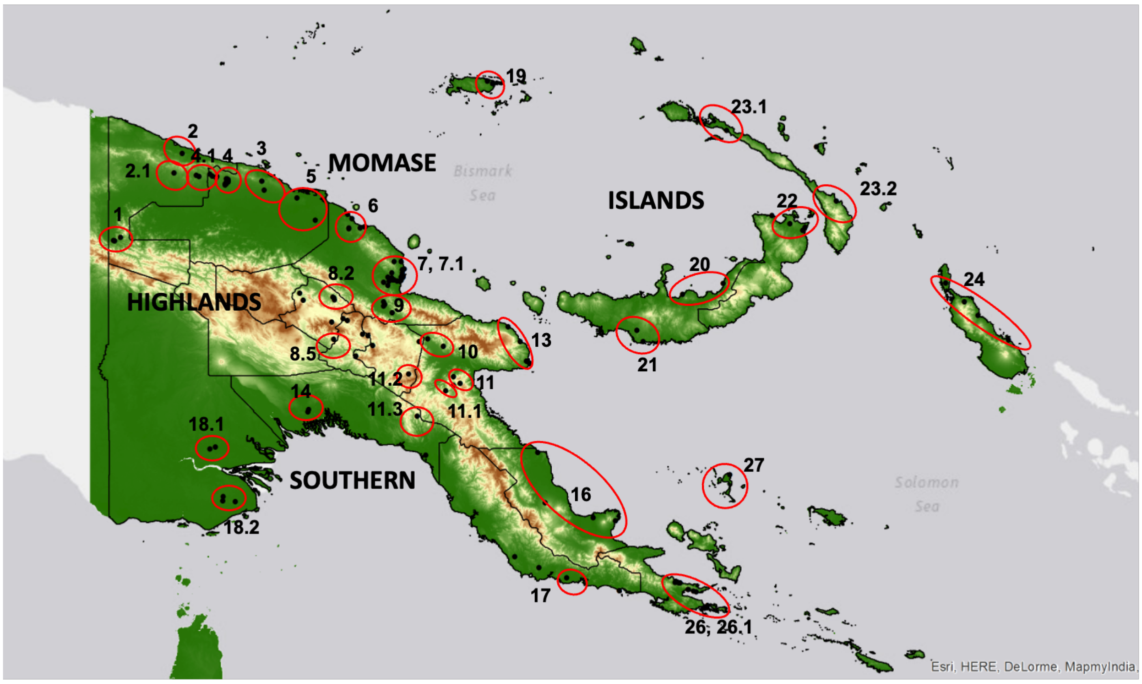


**B.**

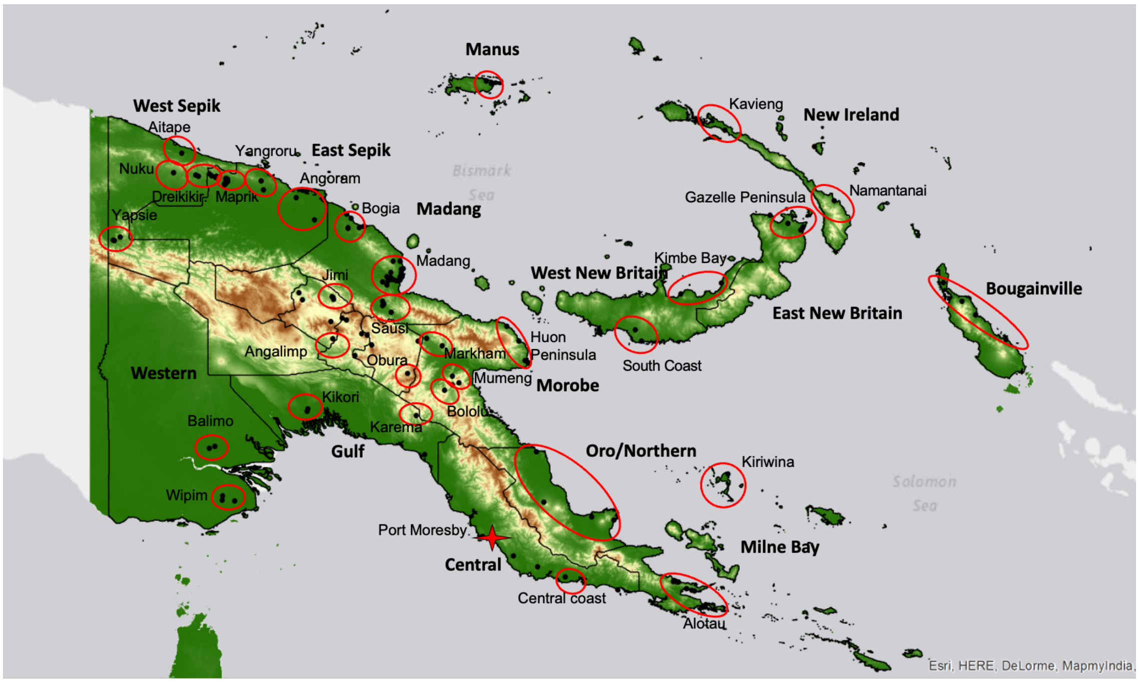


**Figure S3. Map of PNG showing sampling locations of the nationwide survey**
Topographical map of PNG showing lowland (green) and highland areas (brown and yellow). Black dots indicate sampled villages, red circles clusters of villages defined as 'geographic areas' (see **Table S3**). Two maps are provided: A) includes the province (bold) and geographic area names (normal text) referred to in this study. B) includes region names (bold) and geographic area codes (normal text). The map was created using QGIS version 2.14.21 [2].

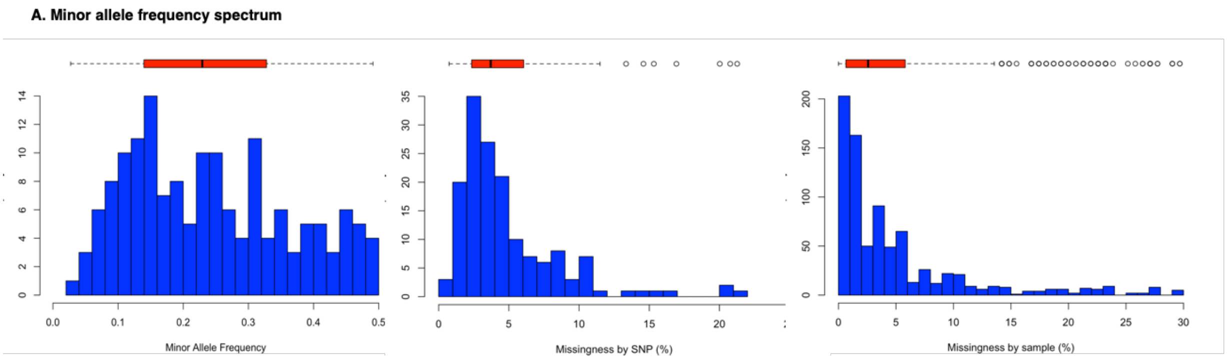


**Figure S4. Quality of the nationwide SNP barcode dataset after filtering.**
Frequency distributions of (A) Minor allele frequencies (MAF), (B) SNP missingness (proportion of SNP calls missing per SNP loci), and (C) sample missingness (proportion of SNP calls missing per samples). The dataset had previously been filtered for SNPs and samples with high missingness (>30% SNP calls missing). The results show a wide range of MAF values demonstrating the informativeness of the barcode for the genotyped populations. For the final dataset used in the analyses (155 SNPs, 733 samples), relatively high SNP and sample coverage (majority of SNPs and samples have less than 10% missingness) was obtained. The boxplots at the top indicate the median (line), interquartile range (box) and 95% confidence intervals (error bars).

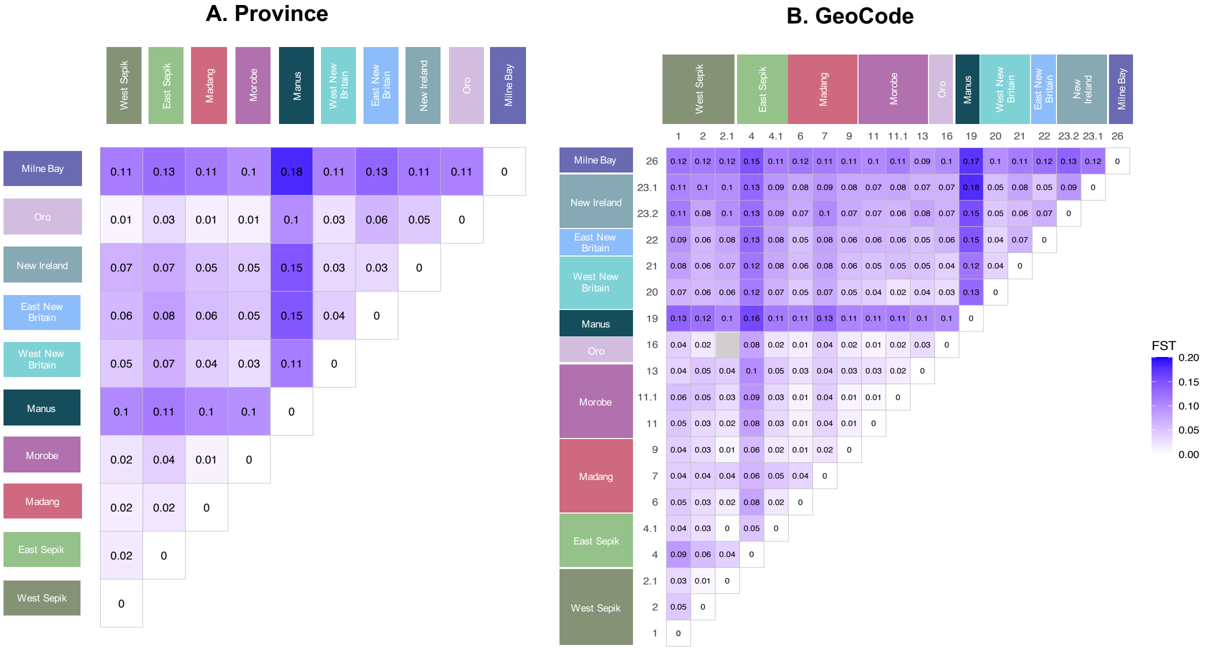


**Figure S5. Genetic differentiation of *P. falciparum* populations from Papua New Guinea.** *Samples were stratified by (A) province and (B) GeoCode and pairwise population level $F_{ST}$ values were estimated to identify geographic population structure. Here, the analysis was restricted to subpopulations with at least ten samples. $F_{ST}$ estimates below zero (which can arise as artefacts due to small sample sizes) are shaded in grey. Colours indicate province as shown in the map in Figure 2A. Geocode names (province: local area) are: 1. West Sepik: Yapsie, 2. West Sepik: Aitape, 2.1 West Sepik: Nuku, 4. East Sepik: Maprik, 4.1. East Sepik: Dreikikir, 6. Madang: Bogia, 7 and 7.1. Madang: Madang, 9. Madang: Sausi+Ramu valley, 11. Morobe: Mumeng, 11.1. Morobe: Bololu, 13. Morobe: Huon Peninsula, 14. Gulf: Kikori, 16. Oro/Northern, 19. Manus, 20. West New Britain: Kimbe Bay, 21. West New Britain: South Coast, 22. East New Britain: Gazelle Peninsula, 23.1. New Ireland: Namantanai, 23.2. New Island: Kavieng, 26 and 26.1. Milne Bay: Alotau. More details on these locations are available in Table S3.*

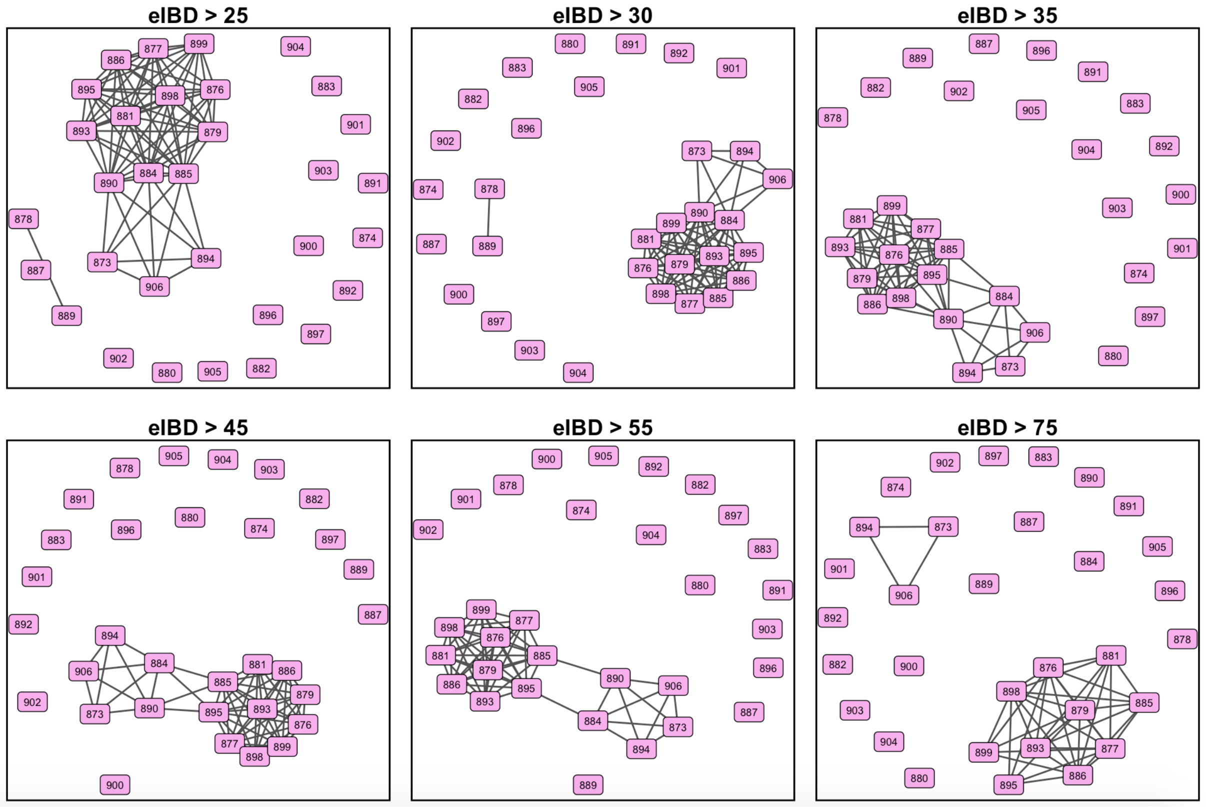


**Figure S6. Relatedness networks of infections from a malaria outbreak amongst migrating workers in Papua New Guinea**

Relatedness networks for infections sampled from the outbreak for a range of eIBD cutoffs, showing the circulation of two parasite clusters with a common genetic background. Below is a table detailing geographic origins of putative imported infections, determined by screening isolates from malaria endemic provinces that exhibited eIBD sharing above 0.25 with infections sampled from the outbreak. eIBD was inferred using IsoRelate [3]. eIBD level indicates the degree of relatedness to the most closely related sample in the nationwide database.

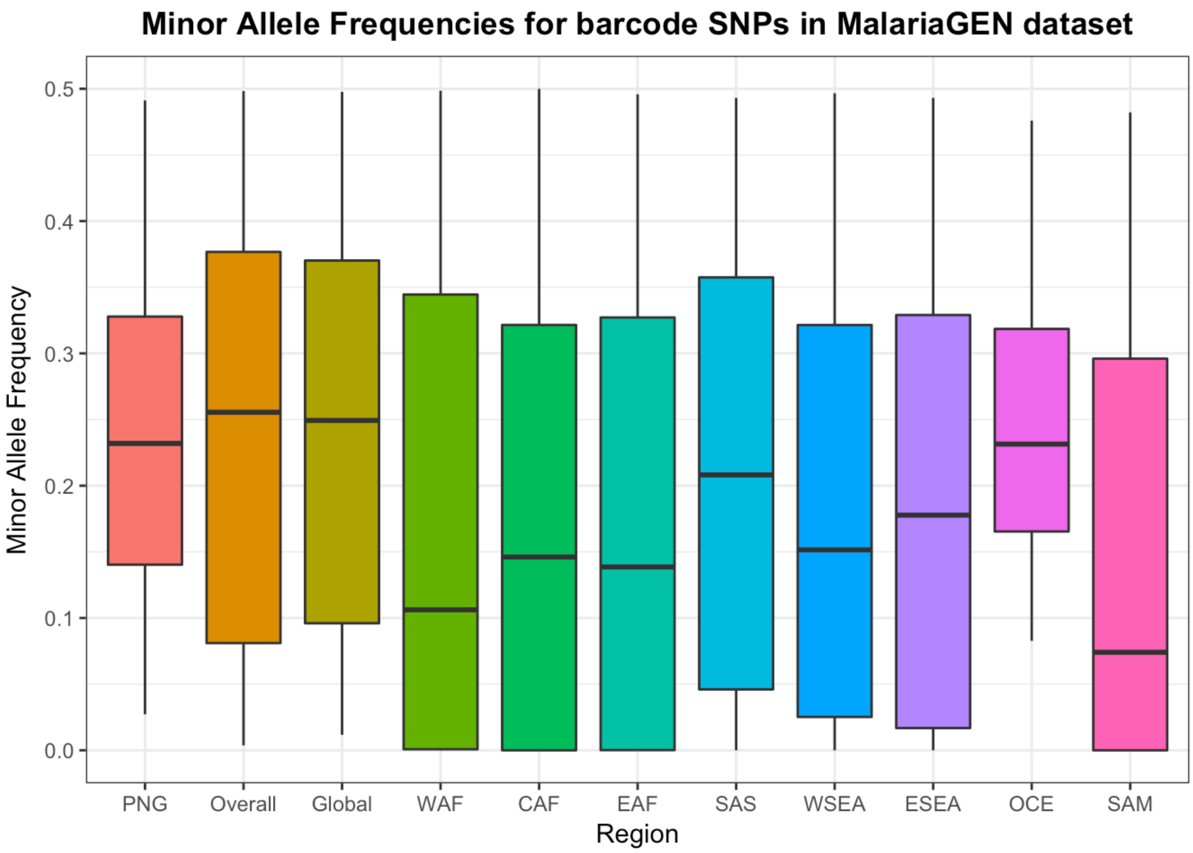


**Figure S7. Minor Allele Frequencies for 155 validated SNPs in different geographic regions.**

Genotypes for the 155 validated local barcode SNPs were extracted from WGS data from 2598 P. falciparum isolates from the publicly available MalariaGEN Pf3k dataset (Version 5.0) [1] and genomes from PNG [1]. Minor allele frequencies were calculated and plotted using boxplots, to show the median (box midline), 75% (boxes) and 95% (lines) interquartile range. Of the 155 validated SNPs, 114 were also polymorphic in African and Asian populations, whilst only 87 were polymorphic amongst South American isolates. Between 90-112 SNPs had a MAF greater than 0.05 in African and Asian populations, and 76 in South America. This shows that despite this barcode being developed for PNG parasites, many of the SNPs will be useful for parasite genomic surveillance in other malaria endemic regions. WAF = West Africa (Benin, Burkina Faso, Cameroon, Ivory Coast, Ghana, Guinea, Mali, Mauritania, Nigeria, Senegal, The Gambia), CAF = Central Africa (Democratic Republic of Congo), EAF = East Africa (Ethiopia, Kenya, Madagascar, Malawi, Tanzania, Uganda, Zambia), SAS =South Asia (Bangladesh), WSEA = Western South East Asia (Myanmar, Western Thailand), ESEA = Eastern South East Asia (Cambodia, Laos, Northeaster Thailand, Viet Nam), Oce = Oceania (Indonesia, PNG), SAM = South America (Colombia, Peru).

### Supporting Tables

**Table S1. Metadata of PNG *P. falciparum* clinical isolates used for whole genome sequencing**
See separate Excel File.

**Table S2. Details of SNP candidates**
See separate Excel File.

**Table S3. Details of geographic catchment areas and sample numbers used for SNP barcoding**
See separate Excel File.

**Table S4. Composition of SNP genotyping reaction mix used in 96.96 Fluidigm Dynamic Array IFC**
(a) Composition of Sample Mix b) Composition of Assay mix for each SNP primers.
See separate Excel File.

**Table S5. Putative origins of imported infections in a malaria outbreak amongst migrating workers in Papua New Guinea**

| | Outbreak Infection | Endemic Infection | eIBD Level | Geographic Origin (Endemic Region) |
|---|---|---|---|---|
| | 874 | 586 | 0.27 | South Coast, West New Britain |
| | 873 | 575 | 0.27 | |
| SAME CLADE | 906 | 575 | 0.27 | |
| | 884 | 481 | 0.25 | Huon Peninsula, Morobe |
| | 902 | 432 | 0.30 | |
| | 882 | 620 | 0.26 | Gazelle Peninsula, East New Britain |
| | 892 | 723 | 0.26 | Kiriwina, Milne Bay |
| | 891 | 33 | 0.29 | Aitape, West Sepik |
